# Planning sEEG implantation using automated lesion detection: retrospective feasibility study

**DOI:** 10.1101/2019.12.08.19013979

**Authors:** Konrad Wagstyl, Sophie Adler, Birgit Pimpel, Aswin Chari, Kiran Seunarine, Sara Lorio, Rachel Thornton, Torsten Baldeweg, Martin Tisdall

**Affiliations:** Wellcome Centre for Human Neuroimaging, University College London, London, UK; Developmental Neurosciences, UCL Great Ormond Street Institute of Child Health, University College London, London, UK; Great Ormond Street Hospital, London, United Kingdom

**Keywords:** epilepsy, neuroimaging, stereoelectroencephalography, paediatric, deep-learning

## Abstract

**Objective:** A retrospective, cross-sectional study to evaluate the feasibility and potential benefits of incorporating deep-learning on structural MRI into planning stereoelectroencephalography (sEEG) implantation in paediatric patients with diagnostically complex drug-resistant epilepsy. This study aims to assess the degree of co-localisation between automated lesion detection and the seizure onset zone (SOZ) as assessed by sEEG.

**Methods:** A neural network classifier was applied to cortical features from MRI data from three cohorts. 1) The network was trained and cross-validated using 34 patients with visible focal cortical dysplasias (FCDs). 2) Specificity was assessed in 20 paediatric healthy controls. 3) Feasibility for incorporation into sEEG implantation plans was evaluated in 38 sEEG patients. Coordinates of sEEG contacts were coregistered with classifier-predicted lesions. sEEG contacts in seizure onset and irritative tissue were identified by clinical neurophysiologists. A distance of <10mm between SOZ contacts and classifier-predicted lesions was considered co-localisation.

**Results:** In patients with radiologically-defined lesions, classifier sensitivity was 74% (25/34 lesions detected). No clusters were detected in the controls (specificity 100%). Of 34 sEEG patients, 21 patients had a focal cortical SOZ. Of these there was co-localisation between classifier output and SOZ contacts in 62%. The algorithm detected 7/8 histopathologically-confirmed FCDs (86%).

**Conclusions:** There was a high degree of co-localisation between automated lesion detection and sEEG. We have created a framework for incorporation of deep-learning based MRI lesion detection into sEEG implantation planning. Our findings demonstrate that automated MRI analysis could be used to plan optimal electrode trajectories.

## Introduction

One third of children with epilepsy are medication-resistant^1^. In children with a focal SOZ, neurosurgical resection can offer seizure freedom in around 70%^2^. Surgical treatment is planned by the multidisciplinary team (MDT) considering results from seizure semiology, neuropsychological, neurodevelopmental, neuropsychiatric evaluation and noninvasive neuroimaging techniques including video-electroencephalography telemetry, MRI, positron-emission tomography and magnetoencephalography. In complex patients, these noninvasive investigations can be inconclusive.

Stereoelectroencephalography (sEEG) can be used to delineate the SOZ in complex patients^3^. In this procedure, implanted depth electrodes directly record brain activity. Currently electrode placement is a clinical decision based on hypotheses generated by the MDT. In half of the patients selected for SEEG, the MRI scan does not show any lesions or shows non-specific abnormalities^4^, limiting the ability to accurately target potential areas of seizure onset.

Using machine-learning, automated lesion detection methods aim to generate putative lesion locations based on structural MRIs^5–7^. We have previously developed a robust and replicated method to identify FCDs^6,8–10^. However, cohorts with confirmed FCDs do not fully capture the complexity and heterogeneity of diagnostically inconclusive patients who present for presurgical evaluation by the MDT.

This retrospective study aimed to create and evaluate a framework for informing and adjusting sEEG electrode planning using automated lesion detection. We trained a classifier to detect focal lesions on patients with MRI-positive FCDs and evaluated it on complex patients who had undergone sEEG. Classifier-identified clusters were co-registered to sEEG electrodes and were assessed for co-localisation the SOZ.

## Methods

### Participants

#### MRI-positive cohort

A retrospective cohort of 34 paediatric patients (mean age = 11.6 years, range = 3.6 - 18.5, female = 20) from Great Ormond Street Hospital (GOSH) was studied, following permission by the hospital ethical review board. Patients were included if they had a radiologically identified focal cortical dysplasia and underwent 3D T1-weighted and FLAIR imaging on the 3T MRI scanner at GOSH. Patients younger than 3 years of age, with MRI scans showing severe motion artefacts (i.e. indistinguishable adjacent gyri due to motion or severe ringing), or without the full protocol described in the following section were excluded.

#### sEEG cohort

All patients who underwent stereo-EEG at GOSH between 2015 and 2018 were identified (n=66, Fig. 1). sEEG patients were excluded for the following reasons: radiological diagnosis of tuberous sclerosis (n=9); hippocampal sclerosis (n=2); vascular/ischaemic lesion (n=4); polymicrogyria (n=1); previous resection (n=12), patients with large MRI detectable lesions where the indication for sEEG was to determine lesion extent (n=4). The final number of sEEG patients was 34 (mean age =11.7 years, range= 3.6-18.5, female=17).

**Figure 1:**
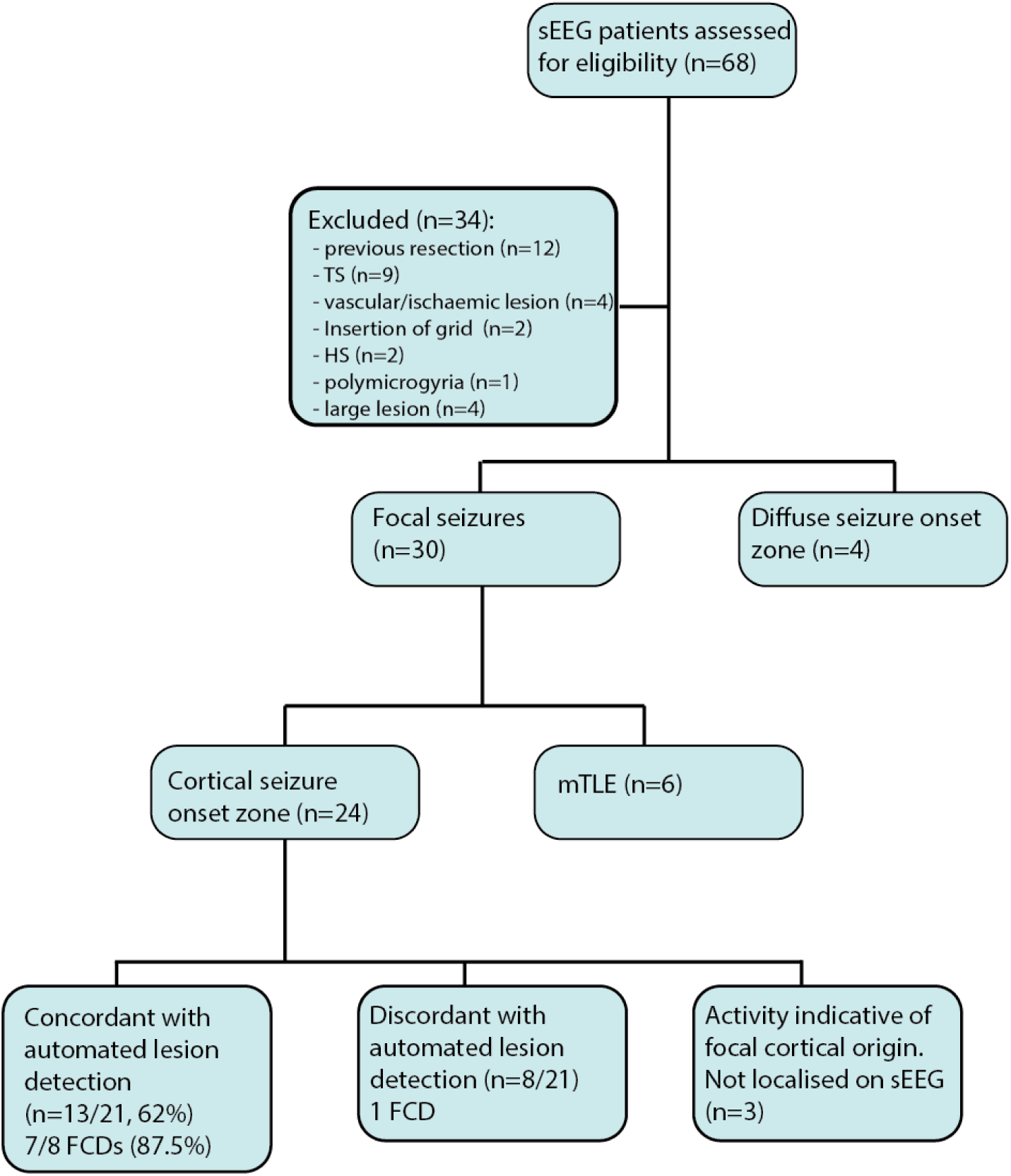
Flowchart of inclusion criteria and MRI / sEEG co-localisation results. Flowchart of inclusion criteria, sEEG results and concordance with automated lesion detection in patients who underwent sEEG.

#### Healthy controls

A control group of 20 term-born children (mean age = 16.8 years, range= 8.4-28.2, female= 14) with no history of any neurological diagnosis were included.

#### MRI acquisition

All patients and controls were scanned on a 3T whole-body MRI system (Magnetom Prisma, Siemens Medical Systems, Germany), using a 20-channel receive head coil and body coil for transmission and 80mT/m magnetic field gradients. Three-dimensional structural T1 weighted (T1w) images and fluid attenuated inversion recovery (FLAIR) images were acquired using the following protocols: magnetisation prepared rapid gradient echo (MPRAGE) (TR=2300ms, TE=2.74ms, FOV=256×256mm, flip angle=8°, voxel size=1×1×1mm^3^) and FLAIR (TR=4000ms, TE=395ms, TI=1800ms, FOV=256×256mm, flip angle=120°, voxel size=0.65×1×0.65mm^3^).

#### MRI post-processing

Post-processing of T1 and FLAIR data followed our previously published automated FCD detection pipeline (https://github.com/kwagstyl/FCDdetection^6^, Fig. 2). In brief:

**Figure 2:**
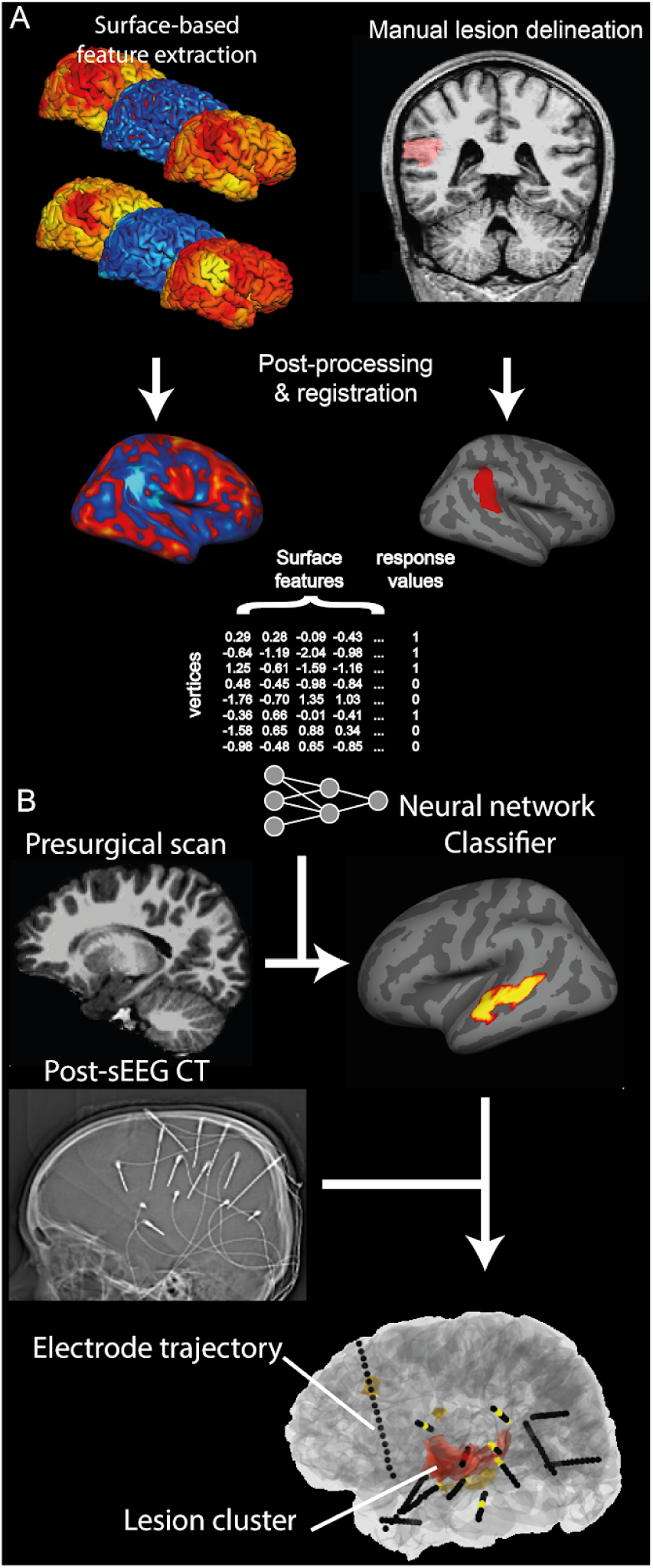
Pipeline for automated lesion detection and co-localisation with sEEG electrodes. A) Surface-based feature extraction, lesion labelling and training of neural network classifier on MRI positive patient cohort. B) Testing of classifier on presurgical MRI of patients undergoing sEEG. Coregistration of classifier output clusters with sEEG electrode implantations extracted from post-surgical CT.

1. Cortical reconstructions were generated using FreeSurfer version 5.3 ^11^.
2. Lesion masks were created for the 34 MRI-positive patients. FCD lesions were identified on T1w and FLAIR images by an experienced paediatric neuroradiologist. 3D binary masks were manually delineated. The lesion masks were first mapped onto the individual surface reconstructions and then to the bilaterally symmetric template (*fsaverage_sym*) ^12^.
3. Measures of morphological/intensity features. The following measures were calculated per vertex across the cortical surface in all participants: 1) cortical thickness, intensity at the grey-white matter contrast, 3) curvature, 4) sulcal depth, 5) intrinsic curvature, and 6) FLAIR signal intensity sampled at 25%, 50% and 75% of the cortical thickness as well as at the grey-white matter boundary and 0.5mm and 1mm subcortically.
4. Smoothing. The following features were smoothed with a 10mm Gaussian kernel: cortical thickness, intensity at the grey-white matter contrast and FLAIR intensities at all cortical and subcortical depths. Intrinsic curvature was smoothed with a 20mm Gaussian kernel to integrate over gyri and sulci.
5. Registration to a bilaterally symmetrical template space. All features were registered to *fsaverage_sym*.
6. Normalisation of features. Features underwent two normalisation procedures. 1)Features were normalised using a within-subject z-scoring, that adjusts for inter-individual differences in the mean and standard deviation. 2) Features were normalised using a between-subject z-scoring, where each participant’s per vertex feature was normalised by the mean and standard deviation in the population of healthy controls. This adjusts for inter-regional differences in the mean and standard deviation.
7. Interhemispheric asymmetry. The right hemisphere vertex values for each feature were subtracted from the left hemisphere values to create a left hemisphere asymmetry map and visa versa for the right hemisphere.
8. Deep learning classification. The Neural Network Toolbox in MATLAB R2018a (The MathWorks, Natick, MA, U.S.A.) was used to create a nonlinear classifier. The structure included 1 hidden layer and the number of nodes in the network was determined through running a principal component analysis (PCA) on the input surface-based features in the control cohort, and using the number of components that explained over 99% of the variance. The neural network was trained using surface based measures from vertices from each MRI-positive patient. The input measures were - normalised cortical thickness, normalised grey-white matter intensity contrast, sulcal depth, mean curvature, the 6 normalised FLAIR intensity samples at different cortical depths and intrinsic curvature as well as the inter-hemispheric asymmetry measures. For training, vertices within the manual lesion masks were extracted as lesional examples while an equal number of randomly selected vertices from the contralateral non-lesional hemisphere were extracted as healthy examples.
9. Clustering and thresholding. Output predictions were grouped into neighbour-connected clusters of vertices with predicted lesion values above a specified threshold. The threshold for the classifier is determined by calculating Youden Index (sensitivity + specificity-100) on the training dataset at a range of values and identifying the optimum threshold values.

#### Evaluation of classifier in lesion positive cohort and controls

To assess the accuracy of the classifier on the lesion-positive cohort, the network was trained using a leave-one-out cross-validation approach, training on 33 lesion-positive subjects and testing on the 34th. Lesions were recorded as being detected if the predicted cluster overlapped with the manually delineated lesion mask. The network was then trained on all 34 patients and tested on the controls to calculate specificity. Any cortical clusters identified by the classifier in controls were recorded as false positives.

#### sEEG

3D electrode trajectories were planned and placed within the patients’ brains using robotic assisted surgery^13^. Pre- and post-surgical CT scans were acquired and coregistered to the presurgical structural MRI using FSL^14^ and visualised using Slicer (www.slicer.org) ^15^. From the post-surgical CT, precise 3D coordinates were calculated for each contact along the depth electrodes (Fig. 2) using an extension for semi-automated electrode contact localiser^16^. sEEG activity was assessed by expert neurophysiologists and individual contacts were classified as being located within the seizure onset zone, irritative zone or not involved in the epileptogenic network. Contacts were considered to be within the seizure onset zone when a typical ictal pattern was seen at onset – stimulation data was also considered where appropriate as is standard practice for sEEG interpretation^17,18^.

#### Comparison between sEEG and MRI clusters

Lesion clusters predicted by the classifier were coregistered from the native MRI surface reconstruction to the surface reconstructions in Slicer space (Fig. 2). The minimum Euclidean distance was calculated from each cluster to each electrode contact.

An automatically identified lesion cluster and sEEG were recorded as co-localised if a lesion cluster was within 10mm of an electrode contact in the seizure onset zone. Neuroimaging processing (KW, SA) and neurophysiology assessment (BP, RT) were carried out independently to avoid bias.

Analysis of how lesion detection would have altered electrode placement In order to estimate the impact on incorporating lesion detection into prospective electrode placement, the number of additional electrodes required to sample potential lesion clusters was calculated for each patient as follows.

Predicted lesion clusters were excluded if:

- They were on the contralateral hemisphere in patients with a unilateral implantation
- Due to an obvious artefact (e.g. motion ringing or skull stripping artefacts)
- Not in the top 3 clusters

If predicted lesion clusters were already within 10mm of an electrode contact, electrodes were classed as ‘adjust’.

The number of remaining clusters which would require an additional electrode or adjustment of an existing nearby electrode were then calculated, using a rule-of-thumb that electrodes within 10mm of an identified cluster could be moved to record from the cluster and those more than 10mm away required an extra electrode to be inserted.

### Data availability

All code to replicate the automated lesion detection analyses and code to compare the automated lesion detection with sEEG depth electrode contacts is freely available from https://github.com/MELDProject. Full results table is also available from https://github.com/MELDProject.

## Results

### Classifier lesion detection results in MRI positive cohort + controls

Out of the 34 patients with visible FCD on MRI, the classifier was able to detect the lesion in 25 (sensitivity = 74%). In the 20 healthy controls no clusters were detected (specificity = 100%).

### sEEG implantation

#### Indication

There were 3 types of indications for sEEG implantation (Table): 1) Discordance: 13 patients were implanted where a lesion had been identified pre-operatively but other data suggested the seizure onset zone may be located elsewhere; 2) 5 patients were implanted as the MRI Imaging was not definitive; 3) 16 patients were implanted as no lesion was identified on MRI (MRI negative).

**Table 1.**

| Pt no. | sEEG indication | sEEG outcome | No. of clusters | Concordance sEEG & automated clusters | Surgery | Histology | Outcome |
| --- | --- | --- | --- | --- | --- | --- | --- |
| 1 | lesion neg | focal | 2 | N | TC & laser | n.a. | SF |
| 2 | lesion neg | focal | 0 | N | Y | non-diag | SF |
| 3 | lesion neg | focal | 3 | Y | Y | non-diag | NSF |
| 4 | lesion neg | focal | 2 | N | N | n.a. | n.a. |
| 5 | lesion neg | focal | 0 | N | N | n.a. | n.a. |
| 6 | lesion neg | focal | 1 | N | Y | FCD 2A | SF |
| 7 | discordance | focal | 4 | Y | Y | FCD 2B | SF |
| 8 | discordance | focal | 7 | Y | Y | non-diag | unknown |
| 9 | discordance | focal | 3 | Y | Y | FCD 2 | SF |
| 10 | discordance | focal | 5 | Y | Y | non-diag | NSF |
| 11 | discordance | focal | 1 | Y | Y | FCD 2B | SF |
| 12 | discordance | focal | 2 | N | Y | other | NSF |
| 13 | discordance | focal | 2 | Y | Y | other | NSF |
| 14 | discordance | focal | 4 | Y | Y | FCD 2B | SF |
| 15 | discordance | focal | 3 | N | Y | non-diag | NSF |
| 16 | not definitive | focal | 1 | Y | Y | FCD 2B | SF |
| 17 | not definitive | focal | 3 | Y | Y | non-diag | SF |
| 18 | not definitive | focal | 2 | Y | Y | FCD IIB | SF |
| 19 | lesion neg | focal | 4 | Y | N | n.a. | n.a. |
| 20 | discordance | focal | 6 | Y | Y | FCD 2A | SF |
| 21 | discordance | focal | 1 | N | TC | n.a. | NSF |
| 22 | lesion neg | mTLE | 1 | n.a. | Y | non-diag | SF |
| 23 | lesion neg | mTLE | 1 | n.a. | Y | non-diag | SF |
| 24 | lesion neg | mTLE | 0 | n.a. | Y | other | NSF |
| 25 | lesion neg | mTLE | 0 | n.a. | Y | non-diag | NSF |
| 26 | lesion neg | mTLE | 2 | n.a. | Y | non-diag | SF |
| 27 | discordance | mTLE | 1 | n.a. | Y | HS | SF |
| 28 | lesion neg | diffuse | 1 | n.a. | N | n.a. | n.a. |
| 29 | lesion neg | diffuse | 4 | n.a. | N | n.a. | n.a. |
| 30 | lesion neg | diffuse | 4 | n.a. | N | n.a. | n.a. |
| 31 | not definitive | diffuse (Rasmussen's) | 1 | n.a. | TC | n.a. | NSF |
| 32 | not definitive | likely focal | 2 | n.a. | TC | n.a. | NSF |
| 33 | discordance | likely focal | 7 | n.a. | N | n.a. | n.a. |
| 34 | lesion neg | likely focal | 6 | n.a. | N | n.a. | n.a. |

*Discordance = discordance between presurgical investigations; mTLE - mesial temporal lobe epilepsy; not definitive = pre-surgical MRI imaging not definitive; lesion neg = lesion negative; non-diag = non-diagnostic; TC = thermocoagulation; Y - yes. N - No (i.e. no co-localisation or no surgery). N.a. Not applicable; FCD - Focal cortical dysplasia; SF - seizure free; NSF - not seizure free.*

#### Outcome of sEEG implantation

In 21 out of 34 patients (62%), a focal cortical epileptogenic seizure onset zone was identified on sEEG (Table). Out of these 21 patients, 16 (76%) underwent subsequent epilepsy surgery and 9 out of 15 (60%) were seizure free at last follow-up (Engel Class 1). In 1 patient, the seizure outcome was unknown. Histology was FCD IIB in 5 patients, FCD IIA in 2, FCD II-unspecified in 1, non-diagnostic in 6 and other in 2 patients. Out of the 8 patients with FCD on histology, all 8 were seizure free at last follow-up (Engel Class 1).

For 7 patients sEEG did not identify focal cortical seizure onset zones. In 3 of these patients, although a focal seizure onset zone was not identified, the pattern of seizure onset was thought to indicate a focal origin where the suspected cortical abnormality was not adequately sampled. 2 of these patients underwent thermocoagulation. In 4 of these patients, seizure onset was described as diffuse, including one patient who has since been diagnosed with Rasmussen encephalitis.

In the final 6 patients, sEEG revealed a mesial temporal lobe seizure onset zone. All 6 patients underwent epilepsy surgery. Histology was non-diagnostic in 4 patients, hippocampal sclerosis in 1 patient and hippocampal gliosis in 1 patient. 4 out of 6 patients (67%) were seizure free at last follow-up (Engel Class 1).

### Comparison of automated lesion detection with sEEG results

Out of the 21 patients in whom a focal cortical epileptogenic seizure onset zone was identified on sEEG, the automatically predicted lesion was co-located with the sEEG determined SOZ in 13 patients (62%, Fig. 1). Out of the 8 patients with histopathologically confirmed FCDs, the predicted lesion was co-located with the sEEG determined SOZ in 7 patients (88%, Fig. 1). Three case studies where there was co-localisation between the predicted lesion and the sEEG ictal contacts are presented in Figure 3.

**Figure 3:**
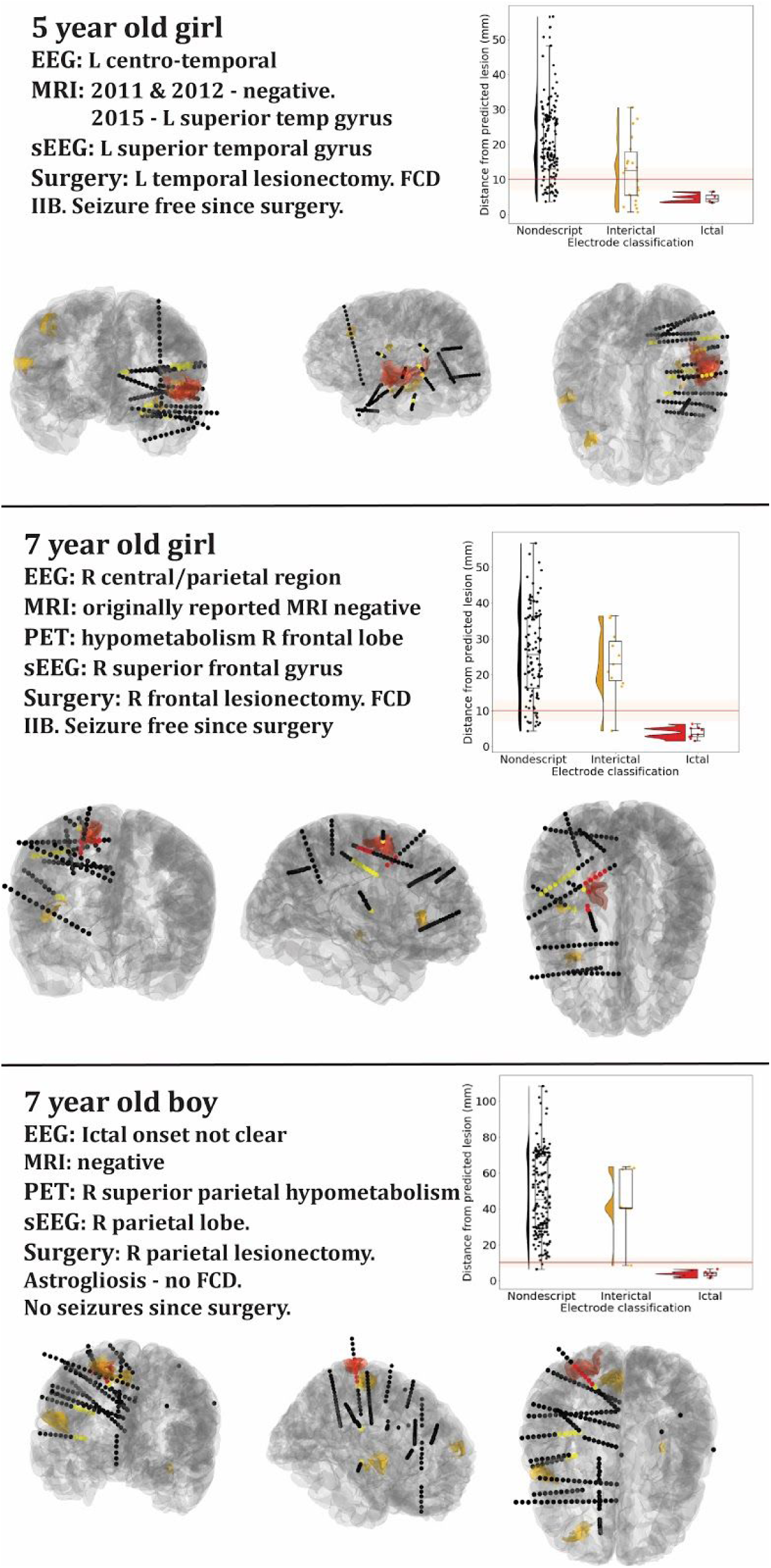
Case studies of three patients. Three example patients where there is co-localisation between ictal contacts and automated lesion detection. Ictal contacts (red contacts) are within 10mm of the automated classifier prediction (red cluster). For each patient are shown: a brief clinical overview (left upper), a plot of distance of the sEEG contacts from the predicted lesion (right upper), and visualisation of the electrode positioning (ictal contacts = red, interictal = yellow, other = black) and automated clusters (red = top cluster, yellow = other clusters, lower panels).

Of the remaining 13 patients, 6 had mesial temporal seizure onset zones identified by sEEG. The automated cortical analysis is unable to explicitly identify mesial temporal (i.e. hippocampal and amygdala abnormalities). However in 2 of these patients, the method did identify abnormalities in the ipsilateral temporal neocortex, consistent with findings that mesial temporal lobe epilepsy is associated with neocortical abnormalities in the ipsilateral temporal lobe ^19^. 2 patients had no neocortical clusters, and 2 had extratemporal neocortical clusters. These are likely false positives.

There were 3 patients in whom the pattern of seizure onset was thought to indicate a focal origin, but where the suspected cortical abnormality was not adequately sampled. In two of these patients, our automated method identified structural cortical abnormalities on the affected hemisphere that were not implanted. It is not possible to retrospectively evaluate whether these are the epileptogenic lesions. A future prospective study could implant these as putative lesion locations.

Over all patients, the minimum number of clusters was 0 and the maximum was 7 (mean±std = 2.53± 1.99).

### Impact on preimplantation planning

Feasibility analysis indicated that 14 extra and 6 adjusted electrodes would be required in order to ensure that the top 3 clusters identified by the classifier were being sampled in all 34 patients. This is an average of 1 extra electrode per 2 patients and 1 adjusted electrode per 6 sEEG patients.

## Discussion

Here we have developed a pipeline for automated detection of FCD and tested the feasibility of incorporating this technology into planning of sEEG trajectories. In the training cohort of patients with radiologically diagnosed FCD our classifier was able to detect 74%, whilst maintaining 100% specificity. In the complex cohort representative of drug resistant epilepsy patients who undergo stereo EEG at a tertiary neurosurgical centre, of the 8 patients who ultimately had histologically confirmed FCD type II, automated lesion detection identified lesional clusters that co-localised with seizure onset zone electrode contacts in 88% (7 patients). Across heterogeneous histopathologies, but with focal seizure onset zones, the automated lesion detection co-localised with seizure onset zone contacts in 62% (13/21 patients). Incorporating automated lesion predictions into implantation strategy would require one additional electrode per two patients. Given the clinical variation in electrode numbers, the potential to identify lesional areas that might be missed and the relatively small increase in bleeding risk of adding an extra electrode to an implantation^20^, this work lays the framework for future prospective studies incorporating these AI technologies into clinical practice.

In recent years there have been considerable advances in the automated detection of focal epileptogenic abnormalities based on structural MRI scans. These approaches use a combination of post-processing and machine learning techniques to automatically delineate structural abnormalities ^5,6,9,21^. However, these studies generally evaluated on cohorts of histopathologically confirmed FCD and healthy controls, which do not reflect the complexity and heterogeneity of patients with medication resistant epilepsy. Therefore it is unclear how these algorithms would perform prospectively and how they might be used to inform clinical decision-making. This study evaluates the feasibility of these techniques on a clinically realistic complex cohort and demonstrates how such approaches could be incorporated in presurgical planning.

The advantage of incorporating these technologies is that it can provide objective lesion hypotheses even in patients in whom the SOZ is difficult to identify and may therefore generate stronger preimplantation hypotheses. In order to test this, a prospective study, where automated lesion detection is incorporated into sEEG implantation planning, is required. Positive outcomes would include increased presurgical confidence, identification of the seizure onset zone in MRI negative patients, improved delineation of cortical lesion boundaries and, ultimately, a potential reduction in the number of required sEEG electrodes. Such developments offer the possibility of improved clinical outcomes and reduced financial burden.

One limitation of this study is that it was retrospective. As there is incomplete sampling of the brain with sEEG, in some patients detected MRI clusters were not close to implanted electrodes. As such it was not possible to determine whether these were false positives or would have exhibited ictal activity, especially in patients where histology was non-diagnostic and patients were not seizure free. However, sEEG is an optimal approach for assessing any new diagnostic technique in focal epilepsy, as it is the only approach for accurate assessment of the ictal onset zone other than post-operative outcome. Thus, this validation problem is shared with any non-invasive technique. A second limitation is the relatively small sample sizes for classifier training and validation. Future studies incorporating MRI data from FCD patients and controls across multiple centres will help to mitigate this^22^.

In conclusion, our study demonstrates the feasibility of incorporating deep learning-based cortical lesion detection from structural MRI into planning of sEEG implantation in patients with suspected focal epilepsy. Additionally we estimated the impact of implanting the extra electrodes required to adequately sample any additional automatically detected structural targets. These analyses lay the foundations for prospective evaluation of automated lesion detection in clinical practice.

## Data Availability

Code and results tables are all available at https://github.com/MELDProject/sEEG

## Acknowledgements

This work and SA was funded by the Rosetrees Trust (A2665). KW was funded by the Wellcome Trust (215901/Z/19/Z). AC is supported by a Great Ormond Street Hospital (GOSH) Children’s Charity Surgeon Scientist Fellowship. This research was supported by the NIHR Great Ormond Street Hospital Biomedical Research Centre. The views expressed are those of the author(s) and not necessarily those of the NHS, the NIHR or the Department of Health. TB is supported by Great Ormond Street Hospital Children’s Charity.

## Notes

**This study was funded by the Rosetrees Trust (A2665)** Dr Wagstyl reports no disclosures. Dr Adler reports no disclosures. Dr Pimpel reports no disclosures. Dr Chari reports no disclosures. Dr Seunarine reports no disclosures. Dr Lorio reports no disclosures. Dr Thornton reports no disclosures.

### Competing Interest Statement

The authors have declared no competing interest.

